# Post-viral mental health sequelae in infected persons associated with COVID-19 and previous epidemics and pandemics: Systematic review and meta-analysis of prevalence estimates

**DOI:** 10.1101/2021.06.29.21259615

**Authors:** Simeon Joel Zürcher, Céline Banzer, Christine Adamus, Anja I. Lehmann, Dirk Richter, Philipp Kerksieck

**Author notes:** **Correspondence:** Simeon J. Zürcher, Center for Psychiatric Rehabilitation, Universitäre Psychiatrische Dienste Bern (UPD), Murtenstrasse 46, 3008 Bern, Switzerland. **Funding:** No funding sources. **Competing interests:** None declared. **Ethical approval:** Not required. **Availability of Data and Materials:** All data to describe the main study characteristics and to calculate prevalence estimates are shown in Table 1 and Supplement. **Author Contributions** SJZ and DR conceived and designed the study. SJZ, CB, CA, and PK conducted the literature search and selection, data extraction, and appraisal of study quality. SJZ conducted the statistical analyses. SJZ, PK, DR and AIL wrote the first draft of the manuscript. All authors contributed to data interpretation, manuscript revisions and approved the final version of the manuscript.

## Abstract

**Aims:** Post-viral mental health problems (MHP) in COVID-19 patients and survivors were anticipated already during early stages of this pandemic. We aimed to synthesize the prevalence of the anxiety, depression, post-traumatic and general distress domain associated with virus epidemics since 2002.

**Methods:** In this systematic review and meta-analysis, we searched PubMed, PsycINFO, and Embase from 2002 until April 14, 2021 for peer-reviewed studies reporting prevalence of MHP in adults with laboratory-confirmed or suspected SARS-CoV-1, H1N1, MERS-CoV, H7N9, Ebolavirus, or SARS-CoV-2 infection. We included studies that assessed post-viral MHP with validated and frequently used scales. A three-level random-effects meta-analysis for dependent sizes was conducted to account for multiple outcome reporting. We pooled MHP across all domains and separately by severity (above mild or moderate-to-severe) and by acute (one month), ongoing (one to three months), and post-illness stages (longer than three months). A meta-regression was conducted to test for moderating effects, particularly for exploring estimate differences between SARS-Cov-2 and previous pandemics and epidemics. PROSPERO registration: CRD42020194535.

**Results:** We identified 59 studies including between 14 to 1002 participants and providing 187 prevalence estimates. MHP, in general, decreased from acute to post-illness from 46□3% to 38□8% and for mild and moderate-to-severe from 22□3% to 18□8%, respectively. We found no evidence of moderating effects except for non-random sampling and H1N1 showing higher prevalence. Pooled MHP differed somewhat between previous pandemics/ epidemics and SARS-CoV-2 but were in a comparable range.

**Conclusions:** MHP prevalence estimates decreased over time but were still on a substantial level at post-illness. Post-viral mental health problems caused by SARS-CoV-2 could have been expected much earlier, given the previous post-viral sequelae.

## 1 Introduction

A large body of evidence from COVID-19 and earlier epidemics such as SARS-CoV-1 and Ebola raised strong concerns regarding acute and long-lasting neurological and psychological problems in infected individuals, now described as Long Covid or Post-acute Covid Syndrome.^1-6^ In COVID-19, this issue was largely underestimated or neglected since public health, and health care priorities focus on safety and survival rather than on mental health care. Even though the importance of acute and long-lasting mental health issues as a consequence of virus infection were highlighted in the context of previous epidemics.^3,7,8^ Although epidemics in the last 20 years like Ebola or COVID-19 differ in many regards like virus characteristics, spread, cultural or socioeconomic environment, they are likely to show important similarities in their impact on mental health in infected individuals or survivors.^3,7,9^ It is known that virus infections like Ebola or Coronaviruses can affect the CNS and cause neuropsychiatric syndromes.^10,11^ For instance, it is assumed that pathophysiological mechanisms including immune response, vascular damage, detrimental effects of critical illness and side effects from treatments may increase the risk for neurodegeneration in COVID-19.^10^ Furthermore, stressors including treatment-related, sociodemographic and environmental circumstances can negatively impact mental health across different epidemics in this population. Infected individuals may suffer from life-threatening complications with uncertain survival or recovery, social isolation, and reduced access to social support, all of which can lead to loneliness and other mental health issues.^12,13^ Affected individuals may face compromised access to health care due to escalating case numbers and overwhelmed health services. Post-illness survivors may need to cope with ongoing symptoms such as reduced physical functioning, fatigue, social and economic issues such as stigmatization with a refusal of services, and reduced working abilities.^1,3,8,14^ Mental health issues may also be aggravated by factors like a history of pre-existing mental health problems.^15^

Within the patient communities affected by post-viral health issues, psychiatric and psychological outcomes are regarded very critically. Similar to communities affected by chronic fatigue syndrome/myalgic encephalomyelitis, these patient initiatives stress biological causal mechanisms over psychological or even social mechanisms.^16,17^ As there are no definitive diagnostic criteria for post-viral health issues such as Long Covid, we have decided to remain neutral in terms of terminology.^18^ Therefore, in the remainder of this paper, we speak of post-viral mental health problems (MHP) rather than symptoms or mental disorders.

Aside from more recent evidence from COVID-19, studies from earlier epidemics are a valuable source of information to inform mental health care. Estimating the magnitude of long-lasting mental and physical problems, including mechanisms and risk factors, is critical to estimate individual, societal and economic costs and facilitate treatment and rehabilitation planning.^19^ The cumulating evidence of potential long-lasting health sequelae associated with Sars-CoV-2 infection is likely to become an important public health issue.

To the best of our knowledge there has been no systematic evidence synthesis published, including patients and survivors of major infections disease epidemics in the last two decades that include all major virus endemics in the last 20 years. In the present meta-analysis, we (1) aimed to overcome this research gap and estimate the pooled prevalence of anxiety, depression, post-traumatic stress, general distress and overall post-viral MHP assessed with validated and widely used scales in suspected and laboratory-confirmed patients and survivors. By ‘post-viral’ we mean MHP including problems that last longer than the acute illness phase (see details in the Methods section). Furthermore, we (2) conducted a meta-regression to investigate potential moderating effects on the overall post-viral MHP prevalence, in particular whether SARS-CoV-2 is different in this regard to previous epidemics and pandemics.

## 2 Material and methods

The study was registered with PROSPERO (CRD42020194535) and is reported in adherence to PRISMA guidelines.^20^ There were some protocol deviations. First, we focused on patients/survivors and excluded original studies on the general population and health-care workers since numerous systematic reviews already covered these populations. Second, we pooled prevalence estimates by domain as well as jointly (see below) rather than by assessment instruments since there was insufficient data to conduct pooling for some instruments. Third, we explicitly looked into differences between SARS-Cov-2 and previous epidemics and pandemics.

### 2.1 Search strategy and selection criteria

We searched PubMed, PsycINFO, Embase and Google Scholar for peer-reviewed studies in the range from January 1, 2002 to April 14, 2021. Reference lists of all eligible studies and topic relevant reviews were screened to identify studies that were potentially missed. We used a broad set of keywords (**Table A.1/A.2**) related to epidemics of interest and assessment instruments. For the latter, we included validated instruments that were used frequently in original studies included based on a similar review conducted in 2020.^7^

Inclusion criteria were: (**a**) peer-reviewed articles using a quantitative methodology; (**b**) published in the languages Dutch, English, French, German or Spanish; (**c**) providing MHP prevalence estimates assessed by any versions of the impact of event scale (IES), the Center for Epidemiologic Studies Depression Scale (CES-D), the Patient Health Questionnaire/Generalized Anxiety Disorder scale (PHQ/GAD), the Hospital Anxiety and Depression Scale (HADS), and the General Health Questionnaire (GHQ); (**d**) adult patients/survivors (≥ 18 years) with suspected or confirmed severe acute respiratory syndrome coronavirus 1 (SARS-CoV-1), swine flu (H1N1), Middle East respiratory syndrome coronavirus (MERS-CoV), avian influenza (H7N9), Ebolavirus, and severe acute respiratory syndrome coronavirus 2 (SARS-CoV-2) infection. Exclusion criteria were: (**a**) subgroups of patients/survivors including psychiatric patients, marginalized individuals, people with chronic physical conditions; (**b**) articles not providing prevalence **(Table A.3)**.

After removing duplicates electronically and manually, two authors (SJZ, PK) identified studies meeting the inclusion criteria based on title and abstracts independently and blinded to each other’s decisions. Any study that met the inclusion criteria was inspected independently and blinded in full-text by two authors (SJZ, CA, CB, PK) for closer inspection. Agreement on full-text eligibility was 90.3% (Cohens Kappa: 0□79 [95% CI 0□69-0□90]). Discrepancies were resolved through discussion.

### 2.2 Data extraction and coding

A standardized form was used to extract study data and quality **(Table A.4)**. Data was extracted and checked by two authors for each included study (SJZ, CA, CB, PK). Variables extracted for descriptive and/or moderator analyses were first authors, year of publication, country, world-region, study design, sampling method, response rate, epidemic, sex ratio, mean/median age, proportion with a history of mental health conditions, proportion in need of intensive care unit, proportion of health care workers, proportion with higher education, treatment received, and months follow-up defined as the time elapsed since treatment or discharge coded as acute (≤ 1 month), ongoing (1-3 months), or post-illness (> 3 months) based on a recently proposed recording system.^21^

The outcome of interest was the prevalence defined as the number of positive classified cases by assessment instrument divided by sample size. We calculated the overall MHP prevalence excluding the GHQ as a not domain-specific measure that assesses new occurring distress-phenomena and carry out normal functions. MHP by domains included anxiety (GAD and HADS scale), depression (PHQ, CES-D, HADS-D scale), post-traumatic stress (IES-Scale), general distress (GHQ scale), and somatization (PHQ-15 Scale). Prevalence values were further stratified by follow-up timepoint and severity defined as at least mild symptoms or at least moderate-to-severe symptoms cut-off by assessment instruments **(Table A.5)**.

### 2.3 Appraisal of the evidence

Quality was appraised independently by SJZ, CA, CB, and PK using eight items of the nine criteria version of the Joanna Briggs Institute Critical Appraisal Checklist for Studies Reporting Prevalence Data.^22^ Each item was rated with yes, no, or unclear and covered sampling frame, sampling/recruitment, sample size, subjects and setting description, coverage, standardized procedures, transparent statistical analyses, and response rate. To the best of our knowledge, there is no published recommendation of weighting and scoring.^22^ We therefore binarized each quality item into yes/no (no or unclear) to calculate the quality achieved in percent (possible range from zero to eight out of a maximum of eight). We classified at least seven points (>87%) as good to excellent, five to six (63-75%) as moderate, less than four (≤50%) as poor. Discrepancies were resolved through discussion. Average inter-rater agreement across all items ranged from 80□0 to 96□7% (Cohens Kappa; 0□75 [95% CI 0□58-0□92] to 0□93 [95% CI 0□83-1□0]).

### 2.4 Data Analysis

We conducted a random-effects meta-analysis for dependent and non-dependent estimates to pool the point prevalence for the overall MHP and for the domain’s anxiety, depression, post-traumatic stress, general distress, and somatization separately. Furthermore, all prevalence estimates were stratified by severity (**Table A.5)** and by follow-up time including acute, ongoing and post-illness stage. In the case of dependency for studies reporting multiple prevalence estimates in the same participants, a three-level mixed-effects model was fitted (taking within-study variation into account).^23^ Freeman-Tukey double-arcsine transformation was used to pool estimates.^24^ We used *I*^*2*^ to determine heterogeneity for analysis in non-dependent prevalence estimates. In dependent estimates, the distribution of total variance (%) attributed to between and within-study variance was estimated.^25^ Sensitivity analyses were performed by the investigation of influential estimates using DFBETAS and Cook’s distance.^26^

We conducted a meta-regression on the overall at least mild and moderate-to-severe MHP prevalence estimates using follow-up timepoint, sex, age, education, history of a mental health condition, health-care workers, duration of hospitalization, intensive care treatment, type of treatment, response rate, sampling method, world region, and epidemic type as moderators. Meta-regression was conducted on arcsine transformed proportions due to better statistical properties as compared to untransformed proportions. All moderators were tested individually while including follow-up timepoint in months. A model including all moderators jointly was not possible due to the substantial missingness of moderators. Predicted prevalence estimates were calculated for each epidemic type to allow a comparison across epidemics. Statistical analyses were conducted in R (version 4.0.3) using metafor.^26^

## 3 Results

### 3.1 Study Characteristics

The systematic search yielded 3304 articles, of which 59 (61 samples) were included (**Figure 1**). The number of individuals ranged from 14 to 1’
s002, with a proportion of females ranging from 22% to 79% and mean/median age from 32-72 years. A total of 187 prevalence rates for mild and moderate-to-severe post-viral MHP prevalence estimates were reported, with one study that assessed samples in different countries^27^ and two that had overlapping samples.^28,29^ Studies covered China (*n*=20), Asia excluding China (*n*=14), Europe (*n*=17), Africa (*n*=6), and America (*n*=3). 42 (71%) studies investigated SARS-CoV-2, eight (14%) SARS-CoV-1, four (7%) MERS-CoV, four (7%) Ebolavirus, and one (2%) H1N1, while no study covered H7N9 (**Table 1**). Time elapsed since treatment/data collection ranged from 0 to about 40 months. Studies rarely reported complete data on all descriptive or moderator variables. While female/male ratio was provided regularly (>98%), other variables such as (history of a psychiatric condition, percent health care workers) were not regularly provided. Overall, 18 studies showed an excellent, 15 studies a moderate and 26 a poor quality on the appraisal scale (**Table 1 and Fig. A.1**).

**Table 1:** Characteristics of included studies reporting mental health problem prevalence estimates in virus disease patients and survivors (*n*=59)

| Author (year) | Country | Virus type | Design | Sampling method<sup>1</sup> | Female (%) | Age<sup>2</sup> | Sample size | Outcomes | Quality<sup>3</sup> |
| --- | --- | --- | --- | --- | --- | --- | --- | --- | --- |
| Akinci and Basar (2021)<sup>30</sup> | Turkey | SARS-CoV-2 | cross-sectional | non-random | 41.3 | 46.3 | 189 | HADS anxiety; HADS depression | 2/8 (25%) |
| Bah et al. (2020)<sup>31</sup> | Sierra Leone | Ebolavirus | cross-sectional | non-random | 50.3 |  | 197 | HADS anxiety; HADS depression | 3/8 (37.5%) |
| Bellan et al. (2021)<sup>32</sup> | Italy | SARS-CoV-2 | longitudinal | random | 40.3 | 61 | 238 | IES-R | 8/8 (100%) |
| Bonazza et al. (2020)<sup>33</sup> | Italy | SARS-CoV-2 | cross-sectional | random | 31.8 | 59 | 184 to 261 | HADS anxiety; HADS depression; IES-R | 6/8 (75%) |
| Chen et al. (2021)<sup>34</sup> | China | SARS-CoV-2 | cross-sectional | non-random | 57.5 | 39.4 | 898 | GAD-7; PHQ-9 | 4/8 (50%) |
| Chen et al. (2020)<sup>35</sup> | China | SARS-CoV-2 | cross-sectional | random | 61.3 | 50 | 31 | GAD-7; PHQ-9 | 2/8 (25%) |
| Cheng et al. (2004)<sup>36</sup> | China | SARS-CoV-1 | cross-sectional | non-random | 66 | 37.1 | 100 | GHQ-28 | 4/8 (50%) |
| Chieffo et al. (2020)<sup>37</sup> | Italy | SARS-CoV-2 | longitudinal | non-random | 44.1 | 54 | 14 to 33 | IES-R | 3/8 (37.5%) |
| D'Cruz et al. (2021)<sup>38</sup> | UK | SARS-CoV-2 | longitudinal | random | 37.8 | 58.7 | 111 to 113 | GAD-7; PHQ-9 | 7/8 (87.5%) |
| Etard et al. (2017)<sup>39</sup> | Guinea | Ebolavirus | longitudinal | random | 54.2 |  | 472 | CES-D | 8/8 (100%) |
| Guo et al. (2020)<sup>40</sup> | China | SARS-CoV-2 | cross-sectional | non-random | 42.7 | 42.5 | 103 | GAD-7; PHQ-9 | 3/8 (37.5%) |
| He et al. (2021)<sup>41</sup> | China | SARS-CoV-2 | cross-sectional | non-random | 50.6 | 56 | 65 | GAD-7; PHQ-9 | 4/8 (50%) |
| Heyns et al. (2021)<sup>42</sup> | Belgium | SARS-CoV-2 | cross-sectional | random | 50.4 | 72 | 47 | HADS anxiety; HADS depression | 7/8 (87.5%) |
| Horn et al. (2020)<sup>43</sup> | France | SARS-CoV-2 | longitudinal | random | 43.9 | 53 | 179 | IES-6 | 7/8 (87.5%) |
| Hu et al. (2020)<sup>44</sup> | China | SARS-CoV-2 | cross-sectional | non-random | 49.4 | 48.8 | 85 | GAD-7; PHQ-9 | 3/8 (37.5%) |
| Islam et al. (2021)<sup>45</sup> | Bangladesh | SARS-CoV-2 | cross-sectional | non-random | 42.1 | 34.7 | 1002 | PHQ-9 | 4/8 (50%) |
| Jeong et al. (2016)<sup>15</sup> | South Korea | MERS-CoV | cross-sectional | non-random | 50 | 52.3 | 36 | GAD-7 | 4/8 (50%) |

**Table 1:** Continued (n=59)
| Author (year) | Country | Virus type | Design | Sampling method<sup>1</sup> | Female (%) | Age<sup>2</sup> | Sample size | Outcomes | Quality<sup>3</sup> |
| --- | --- | --- | --- | --- | --- | --- | --- | --- | --- |
| Jeong et al. (2020)<sup>46</sup> | South Korea | SARS-CoV-2 | longitudinal | non-random | 60.3 | 37.8 | 126 | HADS anxiety; HADS depression | 4/8 (50%) |
| Ju et al. (2021)<sup>47</sup> | China | SARS-CoV-2 | longitudinal | random | 46.3 | 39 | 95 | GAD-7; PHQ-9 | 5/8 (62.5%) |
| Kandeger et al. (2020)<sup>48</sup> | Turkey | SARS-CoV-2 | cross-sectional | random | 44 | 36.7 | 84 | HADS anxiety; HADS depression | 5/8 (62.5%) |
| Kang et al. (2021)<sup>49</sup> | South Korea | SARS-CoV-2 | cross-sectional | random | 52.3 |  | 107 | GAD-7; PHQ-9 | 6/8 (75%) |
| Keita et al. (2017)<sup>50</sup> | Guinea | Ebolavirus | longitudinal | random | 53.9 | 31.6 | 256 | CES-D | 7/8 (87.5%) |
| Kim et al. (2018)<sup>51</sup> | South Korea | MERS-CoV | cross-sectional | random | 63 | 41.1 | 27 | PHQ-9 | 7/8 (87.5%) |
| Kim et al. (2020)<sup>52</sup> | South Korea | SARS-CoV-2 | intervention | random |  | 45 | 33 | HADS anxiety; HADS depression | 5/8 (62.5%) |
| Kong et al. (2020)<sup>53</sup> | China | SARS-CoV-2 | intervention | random | 51.4 | 50 | 144 | HADS anxiety; HADS depression | 4/8 (50%) |
| Kwek et al. (2006)<sup>54</sup> | Singapore | SARS-CoV-1 | cross-sectional | random | 79.4 | 34.8 | 63 | HADS anxiety; HADS depression; IES | 4/8 (50%) |
| Lam et al. (2009)<sup>55</sup> | China | SARS-CoV-1 | cross-sectional | random | 70.4 | 43.3 | 170 | HADS totalscale | 4/8 (50%) |
| Lee et al. (2007)<sup>56</sup> | China | SARS-CoV-1 | cross-sectional | non-random | 63.5 |  | 96 | GHQ-12 | 3/8 (37.5%) |
| Lee et al. (2019)<sup>57</sup> | South Korea | MERS-CoV | longitudinal | random | 38.5 | 49.7 | 52 | IES-R; PHQ-9 | 7/8 (87.5%) |
| Li et al. (2020)<sup>58</sup> | China | SARS-CoV-2 | cross-sectional | non-random | 45.5 | 51.4 | 99 | HADS anxiety; HADS depression | 2/8 (25%) |
| Luyt et al. (2012)<sup>59</sup> | France | H1N1 | longitudinal | random | 51.4 | 39.9 | 37 | HADS anxiety; HADS depression; IES | 7/8 (87.5%) |
| Ma et al. (2020)<sup>12</sup> | China | SARS-CoV-2 | cross-sectional | non-random | 51.9 | 50.4 | 770 | PHQ-9 | 5/8 (62.5%) |
| Mak et al. (2009)<sup>60</sup> | China | SARS-CoV-1 | longitudinal | random | 62.2 | 41.1 | 90 | HADS anxiety; HADS depression | 8/8 (100%) |
| Martillo et al. (2021)<sup>61</sup> | USA | SARS-CoV-2 | longitudinal | random | 22.2 | 53.9 | 42 | PHQ-9 | 6/8 (75%) |

**Table 1:** Continued (n=59)
| Author (year) | Country | Virus type | Design | Sampling method<sup>1</sup> | Female (%) | Age<sup>2</sup> | Sample size | Outcomes | Quality<sup>3</sup> |
| --- | --- | --- | --- | --- | --- | --- | --- | --- | --- |
| Mazza et al. (2020)<sup>62</sup> | Italy | SARS-CoV-2 | cross-sectional | non-random | 34.3 | 57.8 | 102 to 300 | IES-R | 4/8 (50%) |
| Mina et al. (2021)<sup>63</sup> | Bangladesh | SARS-CoV-2 | cross-sectional | non-random | 28 | 39.4 | 145 | GAD-7; PHQ-9 | 2/8 (25%) |
| Morin et al. (2021)<sup>64</sup> | France | SARS-CoV-2 | cross-sectional | random | 38.4 | 56.9 | 169 | HADS anxiety | 8/8 (100%) |
| Mowla et al. (2021)<sup>65</sup> | Iran | SARS-CoV-2 | cross-sectional | random | 35 | 67.4 | 69 | GHQ-28 | 6/8 (75%) |
| Olanipekun et al. (2021)<sup>66</sup> | USA | SARS-CoV-2 | cross-sectional | random | 35.6 | 52.5 | 73 | PHQ-9 | 7/8 (87.5%) |
| Park et al. (2020)<sup>67</sup> | South Korea | MERS-CoV | longitudinal | random | 38.1 | 49.2 | 63 | GAD-7; IES-R; PHQ-9 | 6/8 (75%) |
| Parker et al. (2021)<sup>68</sup> | USA | SARS-CoV-2 | longitudinal | random | 36 | 59 | 58 | HADS anxiety; HADS depression | 6/8 (75%) |
| Paz et al. (2020)<sup>69</sup> | Ecuador | SARS-CoV-2 | cross-sectional | non-random | 51.9 | 37 | 759 | GAD-7; PHQ-9 | 4/8 (50%) |
| Poyraz et al. (2021)<sup>70</sup> | Turkey | SARS-CoV-2 | cross-sectional | random | 49.8 | 39.7 | 284 | HADS anxiety; HADS depression; IES-R | 5/8 (62.5%) |
| Raman et al. (2021)<sup>71</sup> | UK | SARS-CoV-2 | longitudinal | random | 41.4 | 55.4 | 57 | GAD-7; PHQ-9 | 7/8 (87.5%) |
| Rass et al. (2021)<sup>72</sup> | Austria | SARS-CoV-2 | longitudinal | random | 39 | 56 | 98 | HADS anxiety; HADS depression | 7/8 (87.5%) |
| Sahan et al. (2021)<sup>73</sup> | Turkey | SARS-CoV-2 | cross-sectional | random | 49.1 | 55 | 281 | HADS anxiety; HADS depression | 8/8 (100%) |
| Samrah et al. (2020)<sup>74</sup> | Jordan | SARS-CoV-2 | cross-sectional | random | 59.1 | 35.8 | 66 | PHQ-9 | 8/8 (100%) |
| Secor et al. (2020)<sup>27</sup> | Guinea, Liberia, Sierra Leone | Ebolavirus | cross-sectional | random | 57.8 |  | 198 to 751 | GAD-7; PHQ-9 | 5/8 (62.5%) |
| Sheng et al. (2005)<sup>75</sup> | China | SARS-CoV-1 | cross-sectional | non-random | 65.7 | 37.6 | 102 | GHQ-28 | 3/8 (37.5%) |
| Speth et al. (2020)<sup>76</sup> | Switzerland | SARS-CoV-2 | cross-sectional | random | 54.4 | 44.6 | 114 | GAD-2; PHQ-2 | 5/8 (62.5%) |
| van den Borst et al. (2020)<sup>77</sup> | Netherlands | SARS-CoV-2 | longitudinal | random | 40.3 | 59 | 124 | HADS anxiety; HADS depression; IES-R | 7/8 (87.5%) |

**Table 1:** Continued (n=59)
| Author (year) | Country | Virus type | Design | Sampling method<sup>1</sup> | Female (%) | Age<sup>2</sup> | Sample size | Outcomes | Quality<sup>3</sup> |
| --- | --- | --- | --- | --- | --- | --- | --- | --- | --- |
| Wang et al. (2021)<sup>78</sup> | China | SARS-CoV-2 | cross-sectional | random | 64.6 |  | 460 | GAD-7; PHQ-15; PHQ-9 | 6/8 (75%) |
| Wu et al. (2005a)<sup>29</sup> | China | SARS-CoV-1 | cross-sectional | random | 56.9 | 41.5 | 195 | HADS anxiety; HADS depression; IES-R | 7/8 (87.5%) |
| Wu et al. (2005b)<sup>28</sup> | China | SARS-CoV-1 | cross-sectional | non-random | 56 | 41.8 | 131 | HADS anxiety; HADS depression; IES-R | 4/8 (50%) |
| Xu et al. (2021)<sup>79</sup> | China | SARS-CoV-2 | cross-sectional | non-random | 43 | 41.7 | 121 | CES-D | 4/8 (50%) |
| Yadav et al. (2021)<sup>80</sup> | India | SARS-CoV-2 | cross-sectional | non-random | 27 | 42.9 | 100 | GAD-7; PHQ-9 | 2/8 (25%) |
| Zarghami et al. (2020)<sup>81</sup> | Iran | SARS-CoV-2 | cross-sectional | random | 61 | 40.3 | 30 to 52 | GAD-7; PHQ-9 | 5/8 (62.5%) |
| Zhang et al. (2020a)<sup>82</sup> | China | SARS-CoV-2 | cross-sectional | non-random | 50 | 42.5 | 30 | GAD-7; PHQ-9 | 4/8 (50%) |
| Zhang et al. (2020b)<sup>83</sup> | China | SARS-CoV-2 | cross-sectional | non-random | 41.6 |  | 296 | HADS anxiety; HADS depression | 4/8 (50%) |

<sup>1</sup> Random sampling (random or complete sampling strategies where all eligible participants were attempted to be included), non-random sampling (i.e., convenience, snow-ball, or unknown sampling strategy). <sup>2</sup> Refers to mean or median age as provided by original studies. <sup>3</sup> No. of quality items that were answered with yes and percent of full quality complied
CES-D=Center for Epidemiological Studies Depression Scale. GAD-2=Generalized Anxiety Disorder Scale - short form. GAD-7=Generalized Anxiety Disorder Scale. GHQ-12=General Health Questionnaire - short form. GHQ-28=General Health Questionnaire. HADS anxiety=Hospital Anxiety and Depression Scale - anxiety subscale. HADS depression=Hospital Anxiety and Depression Scale - depression subscale. HADS totalscale=Hospital Anxiety and Depression Scale - total-scale. IES-6=Impact of Event Scale - short form. IES-R=Impact of Event Scale-Revised. IES=Impact of Event Scale. PHQ-15=Patient Health Questionnaire somatisation module. PHQ-2=Patient Health Questionnaire Depression module – short form. PHQ-9=Patient Health Questionnaire Depression module.

**Figure 1:**
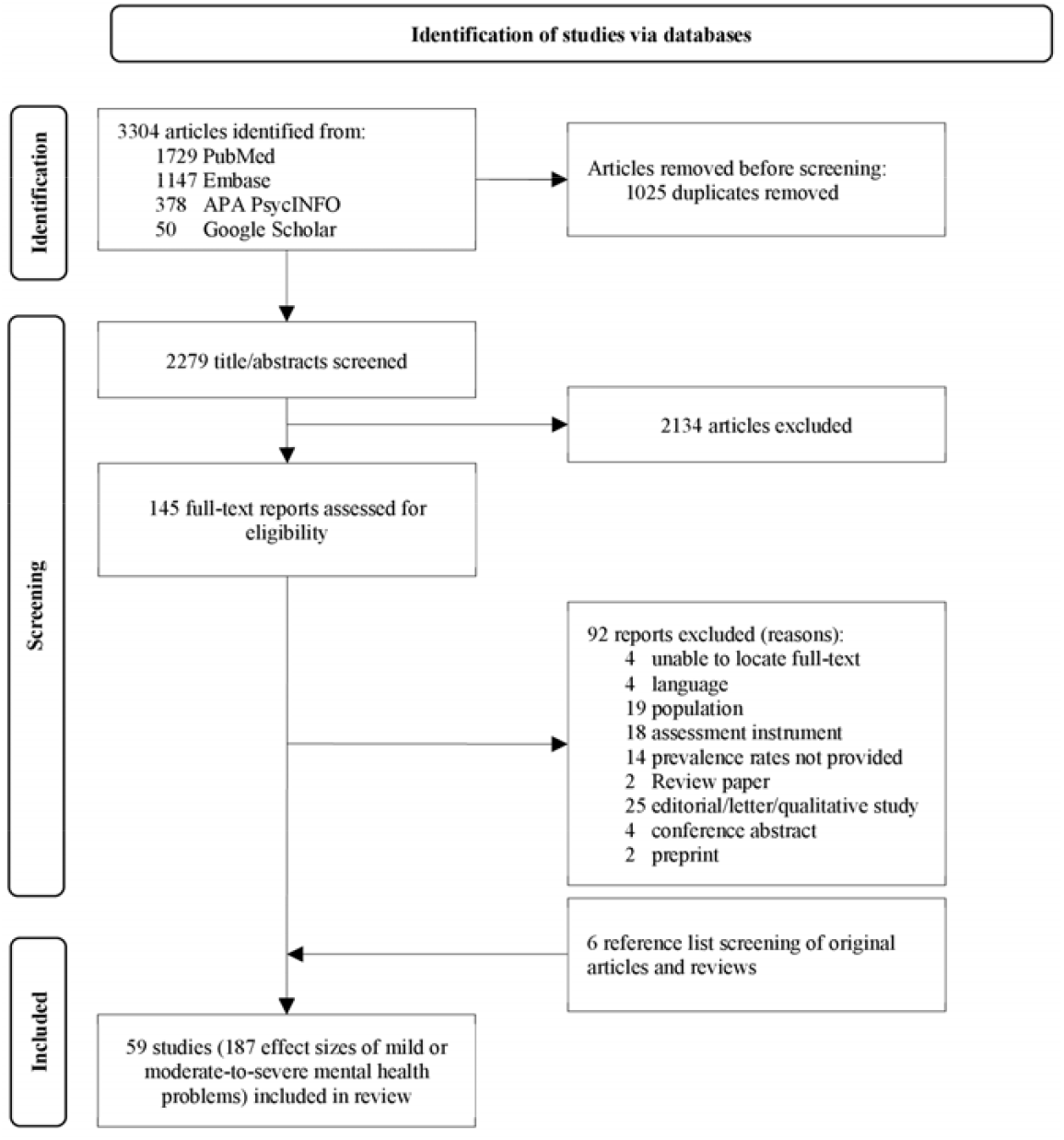
PRISMA Flow chart of included studies reporting on mental health problems in virus disease patients and survivors

### 3.2 Pooled prevalence estimates of mental health problems

Pooled prevalence of mild or moderate-to-severe post-viral MHP including all domains jointly and separately at acute, ongoing and post-illness follow-up timepoint are shown in **Figure 2** and **Table 2**. Original studies contributing to summary estimates are shown in the supplementary material (**Table A.6**).

**Table 2:** Pooled prevalence estimates of at least mild or moderate-to-severe mental health problems including all domains jointly and separately at acute, ongoing, and post-illness stage

| Severity and Domain <sup>1</sup><br>Timepoint of assessment | Nr. effect<br>sizes | Pooled prevalence<br>(95% CI) | p-value (Q) <sup>2</sup> | % between study<br>heterogeneity | % within study<br>variance <sup>3</sup> |
| --- | --- | --- | --- | --- | --- |
| **Mild mental health problems (all domains)** |  |  |  |  |  |
| acute | 55 | 46.3 (39.0-53.8) | <0.0001 | 84.1 | 12.2 |
| ongoing | 11 | 35.5 (18.7-54.3) | <0.0001 | 85.6 | 9.8 |
| post-illness | 18 | 38.8 (33.6-44.1) | <0.0001 | 0.0 | 88.4 |
| **Moderate mental health problems (all domains)** |  |  |  |  |  |
| acute | 60 | 22.3 (17.3-27.8) | <0.0001 | 83.1 | 13.0 |
| ongoing | 22 | 17.3 (10.7-25.1) | <0.0001 | 53.9 | 38.8 |
| post-illness | 17 | 18.8 (13.4-25.0) | <0.0001 | 33.2 | 57.6 |
| **Mild anxiety** |  |  |  |  |  |
| acute | 22 | 44.7 (34.0-55.6) | <0.0001 | 96.3 | 0.0 |
| ongoing | 4 | 28.3 (20.0-37.5) | 0.0188 | 73.6 (*I*<sup>2</sup>) | NA |
| post-illness | 7 | 33.5 (24.8-42.7) | <0.0001 | 0.0 | 89.3 |
| **Moderate anxiety** |  |  |  |  |  |
| acute | 26 | 19.2 (13.5-25.7) | <0.0001 | 95.2 | 0.0 |
| ongoing | 8 | 14.1 (11.1-17.5) | 0.0365 | 55.0 (*I*<sup>2</sup>) | NA |
| post-illness | 6 | 12.1 (5.3-21.1) | <0.0001 | 41.0 | 45.8 |
| **Mild depression** |  |  |  |  |  |
| acute | 30 | 45.3 (38.0-52.7) | <0.0001 | 91.3 | 4.5 |
| ongoing | 5 | 30.8 (11.7-54.1) | <0.0001 | 96.4 (*I*<sup>2</sup>) | NA |
| post-illness | 6 | 43.5 (34.1-53.1) | <0.0001 | 0.0 | 86.6 |
| **Moderate depression** |  |  |  |  |  |
| acute | 26 | 21.9 (15.8-28.6) | <0.0001 | 96.1 | 0.0 |
| ongoing | 9 | 16.0 (8.6-25.2) | <0.0001 | 93.3 (*I*<sup>2</sup>) | NA |
| post-illness | 9 | 18.2 (13.8-23.0) | <0.0001 | 0.0 | 80.3 |
| **Mild post-traumatic stress** |  |  |  |  |  |
| acute | 2 | 62.4 (23.6-93.8) | <0.0001 | 94.8 (*I*<sup>2</sup>) | NA |
| ongoing | 2 | 43.2 (38.0-48.5) | 0.7398 | 0.0 (*I*<sup>2</sup>) | NA |
| post-illness | 4 | 38.6 (24.0-54.3) | 0.2022 | 0.0 | 36.6 |
| **Moderate post-traumatic stress** |  |  |  |  |  |
| acute | 7 | 25.3 (7.5-48.9) | <0.0001 | 94.2 | 2.7 |
| ongoing | 5 | 20.9 (9.2-35.7) | <0.0001 | 95.1 (*I*<sup>2</sup>) | NA |
| post-illness | 2 | 39.1 (25.7-53.2) | 0.3641 | 0.0 (*I*<sup>2</sup>) | NA |
| **Moderate general distress** |  |  |  |  |  |
| acute (NA) |  |  |  |  |  |
| ongoing | 2 | 66.3 (59.6-72.7) | 0.6235 | 0.0 (*I*<sup>2</sup>) | NA |
| post-illness (NA) |  |  |  |  |  |

<sup>1</sup> Shown are pooled effect sizes where at least two studies by domain and timepoint were available. Otherwise indicated with NA. <sup>2</sup> p-value from test of heterogeneity (Q-test). <sup>3</sup> Within study variance displays the amount of within study variances attributed to dependent effect sizes where a three-level meta-analysis was calculated

**Figure 2:**
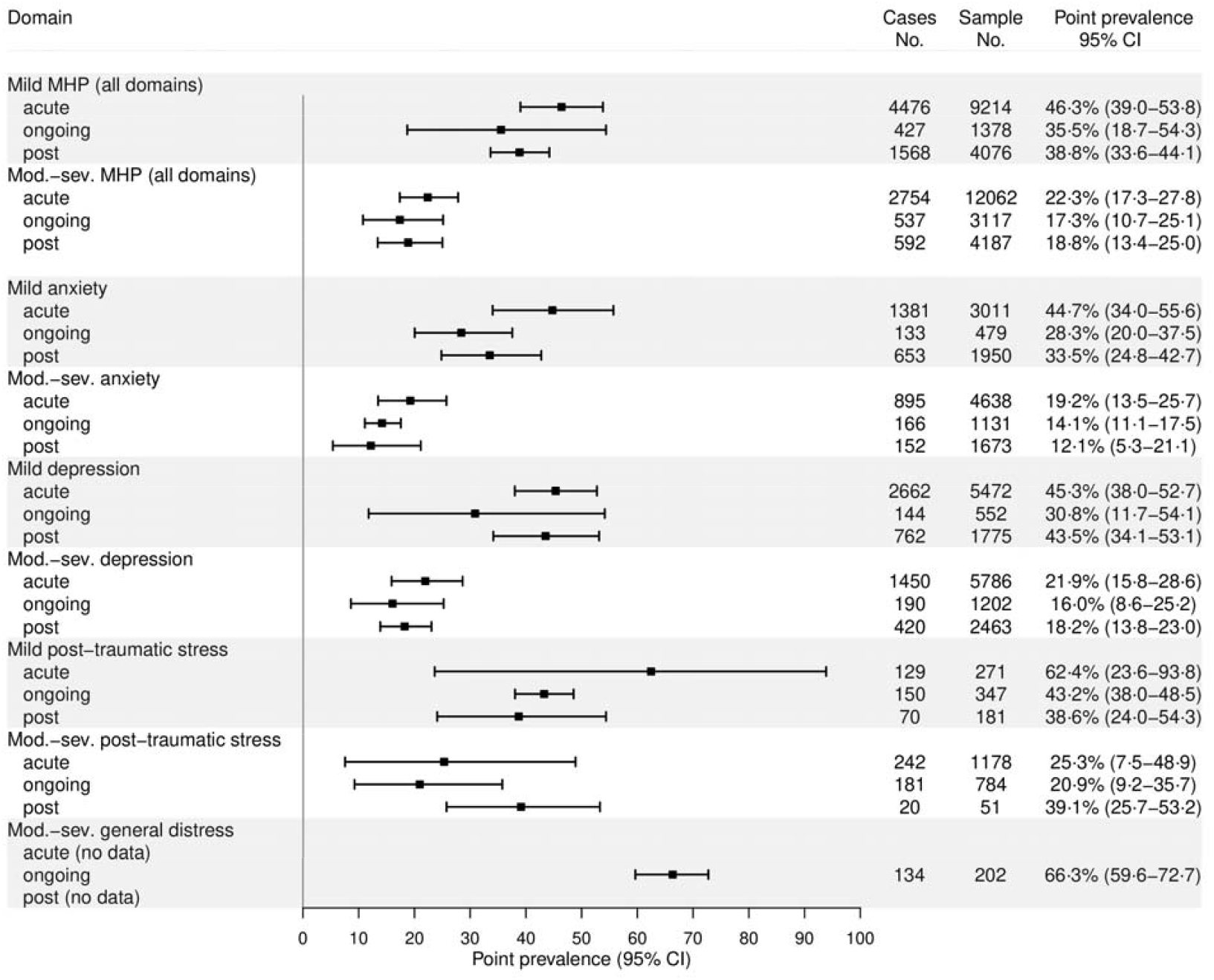
MHP=mental health problems, Mod.-sev.=moderate-to-severe **Caption:** Meta-analysis was conducted only where at least two effect sizes were available. The analysis was conducted with a random effects meta-analysis in the case of independent effect sizes or a three-level random effects model in the case of dependent effect sizes. We used double arcsine transformation for variance stabilization. Displayed are the back-transformed estimates in percent.

Overall mild MHP prevalence was highest in the acute stage with 46□3% and lower at the ongoing and post-illness stage with 35.5% and 38.8%, respectively. Moderate-to-severe MHP estimates were 22.3%, 17.3% and 18.8% for the acute, ongoing, and post-illness stage, respectively. Comparing prevalence estimates between time-points, showed a significant decrease in at least mild and moderate-to-severe MHP between acute and post-illness stage. We found no significant difference between the estimate comparing the acute and ongoing stage (**Table A.7)**.

Although not uniformly, domain-specific estimates for at least mild or moderate-to-severe MHP showed a lower prevalence at post-illness stage as compared to acute stage (**Figure 2** and **Table 2**). For instance, moderate-to-severe anxiety was 19□2% at acute, 14□1% for ongoing, and 12□1% for post-illness. Pooling estimates for somatization, and general distress (acute, post-illness), was not possible since only single prevalence estimates were available (**Table A.6**). To emphasize, the estimates for mild or moderate post-traumatic stress were based on very little data, while the majority of estimators showed substantial heterogeneity with a significant part of variance that could be attributed to between and/or within-study heterogeneity (**Table 2**). Sensitivity analyses did not change the interpretation.

### 3.3 Moderators of mental health problems

The meta-regression to test moderating effects on mild and moderate-to-severe overall MHP prevalence including follow-up time in months is shown in **Table A.7**. We found no evidence of moderating effects for sex, age, education, history of a psychologic condition, health care workers, duration of hospitalization, treatment in ICU, type of treatment, response rate, and world-region. In contrast, we found some evidence for lower mild MHP prevalence in random vs. non-random sampling methods and a higher prevalence of moderate-to-severe MHP in H1N1 vs. SARS-CoV-2. However, the contrast between H1N1 and SARS-CoV-2 should be interpreted cautiously since it is based on very little data. **Figure 3**. shows the predicted moderate-to-severe MHP estimates for the different epidemics (excluding H1N1 since too little data) across the different follow-up timepoints. Generally, estimates show large confidence intervals and seem to differ more in the acute and ongoing phase but lie in a more similar range in the post-illness phase across epidemics.

**Figure 3:**
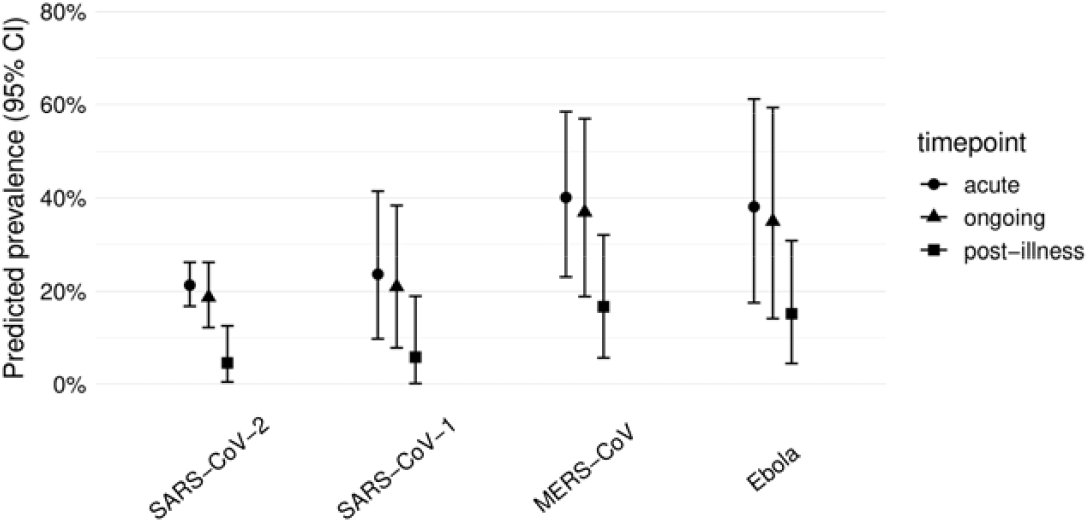
Predicted moderate-to-severe mental health problem prevalence by epidemic/pandemic at acute, ongoing, and post-illness stage **Caption:** Shown are predicted prevalence estimates from the meta-regression for each of the included epidemics at acute, ongoing, and post-illness stage.

## 4 Discussion

A systematic review and meta-analysis was conducted that explored the prevalence estimates of acute, ongoing and post-acute mental health sequelae after infection with SARS-1, MERS, Ebola, H1N1 or SARS-Cov-2 viruses. We included 59 studies providing a total of 187 prevalence estimates in the analysis. A high prevalence of overall and domain-specific MHP in the acute, ongoing and the post-illness stage was identified. Acute infections were associated with higher prevalence estimates of MHP than post-illness. This, however, was not found uniformly across all time-points and mental health problem severity groups.

The overall picture suggests that any mild and moderate-to-severe psychological conditions will be experienced by 39% and 19% of infection survivors for longer than 12 weeks. Likewise, anxiety, depression, and post-traumatic stress, and general distress were substantial beyond the acute phase. Combined, these results show that a considerable proportion of infection survivors will suffer from mental health problems severely for a longer time.

Our results support meta-studies on earlier epidemic outbreaks. A meta-analysis with Chinese publications on SARS-1 found a decrease of mental health problems over time but reported at 12 months post-hospital discharge a level of average distress above population norms.^84^ A systematic review on post-Ebola virus disease studies gathered publications that found considerable depression prevalence and other psychological sequelae in Ebola survivors.^85^

Results reported in our study are based on studies covering multiple epidemics and provide additional information to earlier meta-analytical results referring to COVID-19. A not yet peer-reviewed study on various long-term sequelae of COVID-19 reported a prevalence of 12% for anxiety and 13% for depression.^86^ It is, however, unclear at what time this was assessed. A meta-analysis on early neurological and neuropsychiatric studies (published until July 2020) reported point prevalence values for anxiety of 15.9% and of 23.0% for depression.^1^ A recent large-scale analysis of more than 200,000 electronic health records from the United States found an overall incidence of anxiety and depression diagnoses of 22% within 180 days after Covid-19.^87^ Our results similarly fall in that range. Taken together, we assume that roughly 20% of infected persons develop considerable mental health problems in the weeks and months after an epidemic viral infection. However, milder mental health conditions are somewhat more prevalent and can potentially have detrimental effects as well.

The moderator analysis and the just reported recent research results have shown that post-illness COVID-19 mental health sequelae are not fundamentally different compared to consequences of earlier virus epidemics. While the information on the post-viral health detriments was available from earlier infection outbreaks, it is somewhat astonishing that the risks of longer-term conditions were overlooked in the early days of the pandemic and that it took quite some time to receive the science and media attention it now has.^88^

The moderator analysis has also shown effects for the time point and the sampling method in particular. This suggests that methodological details and study quality are crucial for assessing the contribution of single studies. In general, and as reported from earlier epidemiological studies on mental health effects of epidemics,^89^ the study quality has to be evaluated as mixed. While about 56% reached a moderate to high study quality, about 44 per cent showed a poor quality. Studies frequently suffered from non-transparency regarding sampling frame, recruitment, sample size and provided only a poor description of the study sample and setting and showed low response rates. Furthermore, at least one third showed an unclear or insufficient coverage of the identified sample. In contrast, studies predominantly assessed MHP in a standardized way and clearly described how prevalence was calculated.

Our findings highlight the importance of mental health interventions generally, but at an acute stage of infection specifically. First, clear information about the disease (e.g., infection rates, quarantine, vaccination) is not only important to address uncertainty and fear but also to improve mental health literacy within the population.^90^ Therefore, public communication should also integrate virus- and pandemic-related mental health issues. Furthermore, multidisciplinary mental health support (including psychiatrists, clinical psychologists, mental health nurses and other professions) should be delivered early stage.^91^ Access to mental health interventions could be supplemented by digital health online and smartphone technologies if face-to-face treatment is limited.^90,92,93^ A clinical screening for psychiatric symptoms would ideally be an integrative element already within the acute stage. Therefore, mental health awareness appears to be an important aspect within primary care and emergency departments in particular. Moreover, social support for impaired individuals should be strengthened. This could be supported by prevention strategies that include community-based collaboration among education and employment services, families and housing, and voluntary work.^90^

### 4.1 Strengths and Limitations

The main strength of this research is that it is a large systematic review and meta-analysis that encompasses the major virus outbreaks within the last 20 years. Moreover, this research differentiates between diagnoses, time-points and moderators that allows a comprehensive view regarding the prevalence values of MHP.

This research also has some limitations. Firstly, most studies within this systematic review and meta-analysis lack representativeness. Often, patients with unstable conditions or those within ICU units, or those not hospitalized or later deceased were not included. This selection bias potentially led to an underestimation/overestimation of the prevalence estimates. Also, language bias may be present due to restricting papers based on their original language. Further, many original studies were generally of poor or moderate quality with incomplete data or a lack of random or complete sampling. This might be due to the urgent need for conducting such studies in a pandemic situation. Secondly, this meta-analysis methodology was limited by the fact that few well-validated instruments (e.g., State-Trait Anxiety Inventory) were not included in this research. The meta-regression should be interpreted cautiously since many studies did not provide data on all moderators. Furthermore, it was not possible to include all hypothetical moderators. Specifically, the physical disease severity, the burden of late physical effects or substance abuse were not covered as this was often not reported within studies.

## 5 Conclusion

In this systematic review and meta-analysis covering major virus epidemics in the last 20 years, we found high prevalence rates of at least mild but also moderate-to-severe mental health problems. Moreover, most mental health problems had a higher prevalence at an acute infection stage compared to a post-illness stage. However, post-viral MHP remained substantial in studies covering individuals’ months after infection. Our findings further underline the importance of the study quality that is not often given within the original studies. Therefore, guidelines advising assessment and reporting acute and post-illness MHP in a standardized way are urgently needed. Overall, this research highlights the fragility of mental health after infection from a pandemic virus. Consequently, it emphasizes the need for the early provision of mental health interventions that follow long-lasting post-viral mental health sequelae, particularly rehabilitation interventions.

## Data Availability

Relevant data is provided in the manuscript.

## Acknowledgements

No acknowledgements

