## Supplement for "Post-viral mental health sequelae in infected persons associated with COVID-19 and previous epidemics and pandemics: Systematic review and meta-analysis of prevalence estimates"

**1. Table A.1:** PubMed search terms 2

**2. Table A.2:** PsycInfo and Embase search terms 2

**3. Table A.3:** Inclusion and exclusion criteria 3

**4. Table A.4:** Variables extracted from included studies 4

**5. Table A.5:** Cut-off values by instruments for case detection to categorize mental health problem domains and severity (at least mild or at least moderate-to-severe severity) 5

**6. Fig. A.1:** Summary of methodological quality appraisal of the studies included in the meta-analysis assessed by an adapted version of the Joanna Briggs Institute Critical Appraisal (JBI) tools for Prevalence Studies 6

**7. Table A.6:** Single and pooled prevalence estimates of studies on mental health problems according to domain, severity and timepoint 8

**8. Table A.7:** Test of moderators on at least mild or moderate-to-severe mental health problems including all domains. All moderators were tested separately adjusted for follow-up time in months. 14

**Table A.1:** PubMed search terms

| | |
|---|---|
| **Population** | Betacoronavirus *OR* Sars* *OR* COVID* *OR* 2019-nCoV *OR* Corona *OR* Coronavirus *OR* "Severe Acute Respiratory" *OR* Mers* *OR* "Middle East Respiratory" *OR* H1N1 *OR* Swine-Origin Influenza *OR* "Avian Influenza" *OR* "Avian Flu" *OR* H5N1 *OR* Ebola* |
| **AND** | |
| **Outcomes** | "impact of event scale" *OR* IES *OR* IES-R *OR* "center for epidemiologic studies depression scale" *OR* CES-D *OR* CESD-R *OR* "patient health questionnaire" *OR* PHQ *OR* PHQ-D *OR* PHQ-2 *OR* PHQ-4 *OR* PHQ-7 *OR* PHQ-9 *OR* PHQ-15 *OR* "Generalized Anxiety Disorder" *OR* GAD-2 *OR* GAD-7 *OR* "hospital anxiety and depression scale" *OR* HADS *OR* "general health questionnaire" *OR* GHQ *OR* GHQ-12 *OR* GHQ-28 *OR* GHQ-30 *OR* CHQ |
| **Filter** | From 2002 to current (April 13, 2021) |

**Table A.2:** PsycInfo and Embase search terms

| | |
|---|---|
| **Population** | Betacoronavirus *or* Sars* *or* COVID* *or* 2019-nCoV *or* Corona *or* Coronavirus *or* "Severe Acute Respiratory" *or* Mers* *or* "Middle East Respiratory" *or* H1N1 *or* Swine-Origin Influenza *or* "Avian Influenza" *or* "Avian Flu" *or* H5N1 *or* Ebola* |
| **AND** | |
| **Outcomes** | "impact of event scale" *or* IES *or* IES-R<br>*or* "center for epidemiologic studies depression scale" *or* CES-D *or* CESD-R *or* "patient health questionnaire" *or* PHQ *or* PHQ-D *or* PHQ-2 *or* PHQ-4 *or* PHQ-7 *or* PHQ-9 *or* PHQ-15 *or* "Generalized Anxiety Disorder" *or* GAD-2 *or* GAD-7 *or* "hospital anxiety and depression scale" *or* HADS *or* "general health questionnaire" *or* GHQ *or* GHQ-12 *or* GHQ-28 *or* GHQ-30 *or* CHQ |
| **Filter** | From 2002 to current (April 13, 2021) |

**Table A.3:** Inclusion and exclusion criteria

| | | |
|---|---|---|
| **Population(s)** | **Include** | **Exclude** |
| | *Patients<br>*Survivors<br>*Individuals with suspected infection<br>*Adults (≥18 yrs.) | *Health care workers<br>*General population<br>*Other/special populations (e.g., physical chronical illness, cancer patients, renal transplantation, pregnant women, people with mental illness)<br>*Quarantined individuals (relevant outcomes, setting and characteristics is not reported for cases that test positive or suspected for a virus infection) |
| **Context** | **Include** | **Exclude** |
| | *Virus epidemics including:<br>**SARS-CoV-1<br>**Swine flu (H1N1)<br>**MERS-CoV<br>**Avian influenza (H7N9)<br>**Ebolavirus<br>**SARS-CoV-2 | *Other epidemics (e.g., HIV/AIDS)<br>*Common influenza<br>*Other context (e.g., natural disasters, occupational studies) |
| **Outcome/Condition** | **Include** | **Exclude** |
| | *Prevalence rates provided based on established cut-off scores (or possibility to calculate it) as measured by any short or long form of the following instruments:<br>**Impact of Event Scale (IES)<br>**The Center for Epidemiological Studies Depression Scale (CES-D)<br>**The Patient Health Questionnaire (PHQ)<br>**Generalized Anxiety Disorder Scale (GAD)<br>**The Hospital Anxiety and Depression Scale (HADS)<br>*The General Health Questionnaire (GHQ) | *No prevalence rates provided/calculation not possible (e.g., only mean scores provided)<br>*Prevalence rates assessed by other instruments (e.g., BAI, DASS)<br>*Prevalence rates not based on established cut-off scores (e.g., data-driven cut-off scores like median grouping)<br>*Provided IES prevalence rates for subscales only |
| **Study design/Type** | **Include** | **Exclude** |
| | *All studies that provide prevalence data:<br>*Clinical trial/experimental (only first measure if mental health intervention took place)<br>*Cross-sectional study<br>*Cohort study<br>*Case control study<br>*Case series | *Qualitative study<br>*Psychometric study/questionnaire validation<br>*Editorial/Letter<br>*Mini-study<br>*Review<br>*Conference abstract<br>*Non-scientific article<br>*Studies with $n$<5 |
| **Other criteria** | **Include** | |
| | *Languages: English, German, Spanish, Dutch or French | |

**Table A.4:** Variables extracted from included studies

| Extracted variable | Label/unit |
|---|---|
| DOI | digital object identifier |
| First author | last name of first author |
| Year of publication | YYYY |
| Title of publication | full title |
| County | country |
| World region | Africa, Amerika, Asia (excluding China), China, Europe |
| Design | cross-sectional, longitudinal (repeated measures design), intervention study |
| Sampling | random or complete sample (attempt to include all eligible participants), non-random or unknown sampling strategy (e.g., convenience/snowball) |
| Response rate | numerator: Number of individuals included<br>denominator: Number of eligible individuals (excluding deceased) |
| Epidemic | severe acute respiratory syndrome coronavirus 1 (SARS-CoV-1), swine flu (H1N1), Middle East respiratory syndrome coronavirus (MERS-CoV), avian influenza (H7N9), Ebolavirus, severe acute respiratory syndrome coronavirus 2 (SARS-CoV-2) |
| Sex | % of female sex |
| Age | mean or median and standard deviation |
| Psychological condition | percent with prior psychiatric condition including a history of psychiatric illness or condition or history of psychiatric visits |
| Duration of hospitalization | mean and standard deviation of hospitalization or treatment |
| Intensive care unit | % of included individuals in need of ICU care |
| Health care workers | % of individuals working in the health care sector |
| Education | % of individuals with higher education (tertiary education, university degree) |
| Treatment received | inpatient, outpatient, miscellaneous (inpatient and outpatient) |
| Time point of assessment | mean or median months since discharge/treatment, further coded as acute (≤1 months), ongoing (1-3 Months), post-illness (>3 months) |
| Instrument | IES, IES-R, IES-6 (short form), CES-D, CESD-R (revised), CES-D (short form), PHQ-D (full version), PHQ-9 (depression module), PHQ-2 (depression module – short form), PHQ-4 (depression and anxiety module – short form), PHQ Stress, PHQ-15 (somatization module), PHQ Panic, GAD-2 (short form), GAD-7, GHQ-12, GHQ-28, HADS anxiety, HADS depression, HADS total scale |
| Cut-off score | cut-off used for case detection |
| Number of cases | detected cases |
| Total number | number assessed (number of valid, non-missing responses) |

**Table A.5:** Cut-off values by instruments for case detection to categorize mental health problem domains and severity (at least mild or at least moderate-to-severe severity)

| Assessment instrument and severity classification | Cut-off value for case classification | Severity and domain |
|---|---|---|
| **General Anxiety Disorder Scale** | | |
| GAD-2 (mild) | ≥1 | ≥ mild anxiety |
| GAD-7 (mild) | ≥5 | ≥ mild anxiety |
| GAD-2 (moderate-to-severe) | ≥3 | moderate-to-severe anxiety |
| GAD-7 (moderate-to-severe) | ≥10 | moderate-to-severe anxiety |
| **Hospital Anxiety and Depression Scale; anxiety subscale** | | |
| HADS-Anxiety (mild) | ≥8 | ≥ mild anxiety |
| HADS-Anxiety (moderate-to-severe) | ≥11 (one study with ≥10) | moderate-to-severe anxiety |
| **Patient Health Questionnaire; Depression** | | |
| PHQ-2 (mild) | ≥1 | ≥ mild depression |
| PHQ-9 (mild) | ≥5 | ≥ mild depression |
| PHQ-2 (moderate-to-severe) | ≥3 | moderate-to-severe depression |
| PHQ-9 (moderate-to-severe) | ≥10 | moderate-to-severe depression |
| **Center for Epidemiologic Studies Depression Scale** | | |
| CES-D (no severity classification, indication of depression) | ≥16 or >17 in males and >23 in females | moderate-to-severe depression |
| **Hospital Anxiety and Depression Scale; depression subscale** | | |
| HADS-depression (mild) | ≥8 | ≥ mild depression |
| HADS-depression (moderate-to-severe) | ≥11 (one study with ≥10) | moderate-to-severe depression |
| **Impact of Event Scale** | | |
| IES (mild) | ≥26 | ≥ mild post-traumatic stress |
| IES-R (mild) | ≥24 | ≥ mild post-traumatic stress |
| IES (moderate-to-severe) | ≥30 | moderate-to-severe post-traumatic stress |
| IES-R (moderate-to-severe) | ≥33 | moderate-to-severe post-traumatic stress |
| IES-R (moderate-to-severe) | >2 (surpassing a mean score of 2 in all subscales) | moderate-to-severe post-traumatic stress |
| IES-6 (moderate-to-severe) | ≥10 | moderate-to-severe post-traumatic stress |
| **General Health Questionnaire** | | |
| GHQ-12 | 3 (0-0-1-1 scoring system) | moderate-to-severe general distress |
| GHQ-28 | 24 (0-1-2-3 Likert scale scoring) | moderate-to-severe general distress |
| GHQ-28 | ≥ 5 (0-0-1-1 scoring system) | moderate-to-severe general distress |
| **Hospital Anxiety and Depression Scale; Total scale** | | |
| HADS-anxiety and depression (total scale) | >16 | ≥ mild anxiety and/or depression |
| **Patient Health Questionnaire; somatization module** | | |
| PHQ-15 somatization (mild) | ≥5 | ≥ mild somatization |
| PHQ-15 somatization (moderate-to-severe) | ≥10 | moderate-to-severe somatization |

**Fig. A.1:** Summary of methodological quality appraisal of the studies included in the meta-analysis assessed by an adapted version of the Joanna Briggs Institute Critical Appraisal (JBI) tools for Prevalence Studies

| | Quality Item | | | | | | | |
|---|---|---|---|---|---|---|---|---|
| **Author & Year** | **Sample frame** | **Sampling method** | **Sample size** | **Subjects setting** | **Coverage** | **Procedures** | **Analyses** | **Response Rate** |
| Akinci and Basar (2021) | ? | ✗ | ✗ | ✗ | ? | ✓ | ✓ | ? |
| Bah et al. (2020) | ✗ | ✗ | ✗ | ✗ | ✓ | ✗ | ✓ | ✓ |
| Bellan et al. (2021) | ✓ | ✓ | ✓ | ✓ | ✓ | ✓ | ✓ | ✓ |
| Bonazza et al. (2020) | ✓ | ✓ | ✓ | ✓ | ? | ✓ | ✓ | ? |
| Chen et al. (2021) | ? | ? | ✓ | ✗ | ✓ | ✓ | ✗ | ✓ |
| Chen et al. (2020) | ? | ? | ✗ | ✗ | ✓ | ✗ | ✓ | ? |
| Cheng et al. (2004) | ? | ? | ✗ | ✗ | ✓ | ✓ | ✓ | ✓ |
| Chieffo et al. (2020) | ? | ? | ✗ | ✓ | ? | ✓ | ✓ | ? |
| D´Cruz et al. (2021) | ✓ | ✓ | ✓ | ✓ | ✓ | ? | ✓ | ✓ |
| Etard et al. (2017) | ✓ | ✓ | ✓ | ✓ | ✓ | ✓ | ✓ | ✓ |
| Guo et al. (2020) | ? | ? | ✗ | ✓ | ✓ | ✓ | ✗ | ? |
| He et al. (2021) | ? | ? | ✗ | ✓ | ✓ | ✓ | ✓ | ✗ |
| Heyns et al. (2021) | ✓ | ✓ | ✓ | ✓ | ✓ | ✓ | ✓ | ✗ |
| Horn et al. (2020) | ✓ | ✓ | ✓ | ✓ | ✓ | ✓ | ✗ | ✓ |
| Hu et al. (2020) | ? | ? | ✗ | ✓ | ✓ | ✗ | ✓ | ✗ |
| Islam et al. (2021) | ✗ | ✗ | ✓ | ✗ | ✗ | ✓ | ✓ | ✓ |
| Jeong et al. (2016) | ✓ | ✗ | ✗ | ✗ | ✓ | ✓ | ✓ | ✗ |
| Jeong et al. (2020) | ? | ? | ✗ | ✓ | ✓ | ✓ | ✓ | ? |
| Ju et al. (2021) | ✓ | ✓ | ✓ | ✓ | ? | ✓ | ? | ✗ |
| Kandeger et al. (2020) | ✓ | ✓ | ✓ | ✗ | ✓ | ✓ | ✗ | ✗ |
| Kang et al. (2020) | ✓ | ✓ | ✓ | ✗ | ✓ | ✓ | ✗ | ✓ |
| Keita et al. (2017) | ✓ | ✓ | ✓ | ✓ | ✓ | ✓ | ✓ | ✗ |
| Kim et al. (2018) | ✓ | ✓ | ✓ | ✓ | ✓ | ✓ | ✓ | ✗ |
| Kim et al. (2020) | ✓ | ✓ | ✓ | ✗ | ? | ✓ | ✓ | ✗ |
| Kong et al. (2020) | ? | ? | ✗ | ✗ | ✓ | ✓ | ✓ | ✓ |
| Kwek et al. (2006) | ? | ✓ | ? | ✓ | ✗ | ? | ✓ | ✓ |
| Lam et al. (2009) | ✓ | ✓ | ✓ | ✗ | ? | ✓ | ✗ | ✗ |
| Lee et al. (2007) | ? | ? | ? | ✗ | ? | ✓ | ✓ | ✓ |
| Lee et al. (2019) | ✓ | ✓ | ✓ | ✓ | ? | ✓ | ✓ | ✓ |
| Li et al. (2020) | ? | ? | ✗ | ✗ | ✓ | ? | ✓ | ✗ |
| Luyt et al. (2012) | ✓ | ✓ | ✓ | ✓ | ✓ | ? | ✓ | ✓ |
| Ma et al. (2020) | ? | ? | ✓ | ? | ✓ | ✓ | ✓ | ✓ |
| Mak et al. (2009) | ✓ | ✓ | ✓ | ✓ | ✓ | ✓ | ✓ | ✓ |
| Martillo et al. (2021) | ✓ | ✓ | ✓ | ✓ | ✗ | ✓ | ✓ | ✗ |
| Mazza et al. (2020) | ? | ? | ? | ✓ | ✓ | ✓ | ✓ | ✗ |
| Mina et al. (2021) | ✗ | ✗ | ✗ | ✗ | ✗ | ✓ | ✓ | ✗ |

**Fig. A.1:** Continued

| | Quality Item | | | | | | | |
|---|---|---|---|---|---|---|---|---|
| **Author & Year** | **Sample frame** | **Sampling method** | **Sample size** | **Subjects setting** | **Coverage** | **Proce-dures** | **Analyses** | **Response Rate** |
| Morin et al. (2021) | ✓ | ✓ | ✓ | ✓ | ✓ | ✓ | ✓ | ✓ |
| Mowla et al. (2021) | ✓ | ✓ | ✓ | ✗ | ✗ | ✓ | ✓ | ✓ |
| Olanipekun et al. (2021) | ✓ | ✓ | ✓ | ✓ | ✗ | ✓ | ✓ | ✓ |
| Park et al. (2020) | ✓ | ✓ | ✓ | ✓ | ✓ | ? | ✓ | ✗ |
| Parker et al. (2021) | ✓ | ✓ | ✓ | ✓ | ✗ | ✓ | ✓ | ✗ |
| Paz et al. (2020) | ✗ | ? | ✓ | ✗ | ✓ | ✓ | ✓ | ✗ |
| Poyraz et al. (2021) | ✓ | ✓ | ✓ | ✓ | ✗ | ✗ | ✓ | ✗ |
| Raman et al. (2021) | ✓ | ✓ | ✓ | ✓ | ✓ | ✓ | ✓ | ✗ |
| Rass et al. (2021) | ✓ | ✓ | ✓ | ✓ | ✓ | ? | ✓ | ✓ |
| Sahan et al. (2021) | ✓ | ✓ | ✓ | ✓ | ✓ | ✓ | ✓ | ✓ |
| Samrah et al. (2020) | ✓ | ✓ | ✓ | ✓ | ✓ | ✓ | ✓ | ✓ |
| Secor et al. (2020) | ✓ | ✓ | ✓ | ✗ | ? | ✓ | ✓ | ? |
| Sheng et al. (2005) | ? | ? | ✗ | ✓ | ✗ | ✓ | ✓ | ? |
| Speth et al. (2020) | ✓ | ✓ | ✓ | ✗ | ✓ | ? | ✓ | ✗ |
| van den Borst et al. (2020) | ✓ | ✓ | ✓ | ✓ | ✓ | ? | ✓ | ✓ |
| Wang et al. (2021) | ✓ | ✓ | ✓ | ✗ | ✗ | ✓ | ✓ | ✓ |
| Wu et al. (2005a) | ✓ | ✓ | ✓ | ✓ | ✓ | ✓ | ✓ | ✗ |
| Wu et al. (2005b) | ? | ? | ? | ✗ | ✓ | ✓ | ✓ | ✓ |
| Xu et al. (2021) | ✓ | ? | ? | ✗ | ✓ | ✓ | ✓ | ? |
| Yadav et al. (2021) | ? | ? | ? | ✓ | ✗ | ? | ✓ | ✗ |
| Zarghami et al. (2020) | ✓ | ✓ | ✓ | ✓ | ? | ✓ | ✗ | ✗ |
| Zhang et al. (2020a) | ? | ✗ | ✗ | ✓ | ✓ | ✓ | ✓ | ? |
| Zhang et al. (2020b) | ✗ | ✗ | ✓ | ✗ | ✓ | ✓ | ✓ | ? |

✓ =yes; ✗ =no; ? =unclear

Sample frame=was the sample frame appropriate to assess the target population? Sampling=were the study participants selected/recruited in an appropriate way? Sample size=was the sample size adequate? Subjects Setting=were the study subjects and setting described in detail? Coverage=was data analysis conducted with sufficient coverage of the identified sample? Procedures=was the condition measured in a standardized, reliable way for all participants? Analyses=was there appropriate statistical analysis? Response rate=was the response rate adequate, and if not, was the low response rate managed appropriately?

**Table A.6:** Single and pooled prevalence estimates of studies on mental health problems according to domain, severity and timepoint

| Severity, Domain, Timepoint<br>First author | Year | Country | Epidemic | No. of cases | Sample size | Pooled prevalence (95% CI) |
|---|---|---|---|---|---|---|
| **At least mild anxiety, acute** | | | | | | |
| Chen et al. | 2020 | China | SARS-CoV-2 | 6 | 31 | 19·4 (7·0-35·4) |
| Guo et al. | 2020 | China | SARS-CoV-2 | 57 | 103 | 55·3 (45·6-64·9) |
| He et al. | 2021 | China | SARS-CoV-2 | 34 | 65 | 52·3 (40·1-64·4) |
| Heyns et al. | 2021 | Belgium | SARS-CoV-2 | 16 | 47 | 34·0 (21·1-48·3) |
| Hu et al. | 2020 | China | SARS-CoV-2 | 33 | 85 | 38·8 (28·7-49·5) |
| Ju et al. | 2021 | China | SARS-CoV-2 | 41 | 95 | 43·2 (33·3-53·3) |
| Ju et al. | 2021 | China | SARS-CoV-2 | 36 | 95 | 37·9 (28·4-47·9) |
| Kang et al. | 2020 | South Korea | SARS-CoV-2 | 16 | 107 | 15·0 (8·8-22·4) |
| Kim et al. | 2020 | South Korea | SARS-CoV-2 | 6 | 33 | 18·2 (6·6-33·4) |
| Kong et al. | 2020 | China | SARS-CoV-2 | 50 | 144 | 34·7 (27·1-42·7) |
| Li et al. | 2020 | China | SARS-CoV-2 | 41 | 99 | 41·4 (31·9-51·3) |
| Mina et al. | 2021 | Bangladesh | SARS-CoV-2 | 92 | 145 | 63·4 (55·4-71·1) |
| Park et al. | 2020 | South Korea | MERS-CoV | 44 | 63 | 69·8 (57·9-80·6) |
| Parker et al. | 2021 | USA | SARS-CoV-2 | 21 | 58 | 36·2 (24·3-49·1) |
| Paz et al. | 2020 | Ecuador | SARS-CoV-2 | 441 | 759 | 58·1 (54·6-61·6) |
| Speth et al. | 2020 | Switzerland | SARS-CoV-2 | 51 | 114 | 44·7 (35·7-54·0) |
| Wang et al. | 2021 | China | SARS-CoV-2 | 213 | 460 | 46·3 (41·8-50·9) |
| Yadav et al. | 2021 | India | SARS-CoV-2 | 67 | 100 | 67·0 (57·4-75·9) |
| Zarghami et al. | 2020 | Iran | SARS-CoV-2 | 7 | 30 | 23·3 (9·7-40·4) |
| Zarghami et al. | 2020 | Iran | SARS-CoV-2 | 17 | 52 | 32·7 (20·5-46·1) |
| Zhang et al. | 2020a | China | SARS-CoV-2 | 30 | 30 | 100·0 (94·3-100·0) |
| Zhang et al. | 2020b | China | SARS-CoV-2 | 62 | 296 | 20·9 (16·5-25·8) |
| **Summary Effect** | | | | **1381** | **3011** | **44·7 (34·0-55·6)** |
| **At least mild anxiety, ongoing** | | | | | | |
| Bonazza et al. | 2020 | Italy | SARS-CoV-2 | 73 | 261 | 28·0 (22·7-33·6) |
| Kwek et al. | 2006 | Singapore | SARS-CoV-1 | 21 | 63 | 33·3 (22·2-45·5) |
| Raman et al. | 2021 | UK | SARS-CoV-2 | 22 | 57 | 38·6 (26·3-51·6) |
| Rass et al. | 2021 | Austria | SARS-CoV-2 | 17 | 98 | 17·3 (10·4-25·5) |
| **Summary Effect** | | | | **133** | **479** | **28·3 (20·0-37·5)** |
| **At least mild anxiety, post-illness** | | | | | | |
| Bah et al. | 2020 | Sierra Leone | Ebolavirus | 49 | 197 | 24·9 (19·1-31·2) |
| Luyt et al. | 2012 | France | H1N1 | 20 | 37 | 54·1 (37·7-69·9) |
| Morin et al. | 2021 | France | SARS-CoV-2 | 53 | 169 | 31·4 (24·6-38·6) |
| Park et al. | 2020 | South Korea | MERS-CoV | 20 | 63 | 31·7 (20·8-43·8) |
| Secor et al. | 2020 | Liberia | Ebolavirus | 74 | 203 | 36·5 (30·0-43·2) |
| Secor et al. | 2020 | Sierra Leone | Ebolavirus | 317 | 751 | 42·2 (38·7-45·8) |
| Secor et al. | 2020 | Guinea | Ebolavirus | 120 | 530 | 22·6 (19·2-26·3) |
| **Summary Effect** | | | | **653** | **1950** | **33·5 (24·8-42·7)** |

**Table A.6:** Continued

| Severity, Domain, Timepoint<br>First author | Year | Country | Epidemic | No. of cases | Sample size | Pooled prevalence (95% CI) |
|---|---|---|---|---|---|---|
| **Moderate-to-severe anxiety, acute** | | | | | | |
| Akinci and Basar | 2021 | Turkey | SARS-CoV-2 | 25 | 189 | 13·2 (8·7-18·5) |
| Chen et al. | 2021 | China | SARS-CoV-2 | 147 | 898 | 16·4 (14·0-18·9) |
| Chen et al. | 2020 | China | SARS-CoV-2 | 3 | 31 | 9·7 (1·3-23·1) |
| Guo et al. | 2020 | China | SARS-CoV-2 | 7 | 103 | 6·8 (2·6-12·6) |
| He et al. | 2021 | China | SARS-CoV-2 | 10 | 65 | 15·4 (7·5-25·3) |
| Hu et al. | 2020 | China | SARS-CoV-2 | 14 | 85 | 16·5 (9·3-25·2) |
| Jeong et al. | 2016 | South Korea | MERS-CoV | 17 | 36 | 47·2 (31·0-63·7) |
| Jeong et al. | 2020 | South Korea | SARS-CoV-2 | 13 | 126 | 10·3 (5·5-16·3) |
| Ju et al. | 2021 | China | SARS-CoV-2 | 16 | 95 | 16·8 (9·9-25·1) |
| Ju et al. | 2021 | China | SARS-CoV-2 | 11 | 95 | 11·6 (5·8-18·9) |
| Kandeger et al. | 2020 | Turkey | SARS-CoV-2 | 16 | 84 | 19·0 (11·3-28·2) |
| Kang et al. | 2020 | South Korea | SARS-CoV-2 | 2 | 107 | 1·9 (0·0-5·6) |
| Kong et al. | 2020 | China | SARS-CoV-2 | 25 | 144 | 17·4 (11·6-24·0) |
| Li et al. | 2020 | China | SARS-CoV-2 | 15 | 99 | 15·2 (8·7-23·0) |
| Mina et al. | 2021 | Bangladesh | SARS-CoV-2 | 38 | 145 | 26·2 (19·3-33·7) |
| Mina et al. | 2021 | Bangladesh | SARS-CoV-2 | 42 | 145 | 29·0 (21·8-36·6) |
| Park et al. | 2020 | South Korea | MERS-CoV | 33 | 63 | 52·4 (40·0-64·7) |
| Parker et al. | 2021 | USA | SARS-CoV-2 | 8 | 58 | 13·8 (5·9-24·0) |
| Paz et al. | 2020 | Ecuador | SARS-CoV-2 | 171 | 759 | 22·5 (19·6-25·6) |
| Sahan et al. | 2021 | Turkey | SARS-CoV-2 | 98 | 281 | 34·9 (29·4-40·6) |
| Speth et al. | 2020 | Switzerland | SARS-CoV-2 | 12 | 114 | 10·5 (5·5-16·9) |
| Wang et al. | 2021 | China | SARS-CoV-2 | 91 | 460 | 19·8 (16·3-23·6) |
| Wu et al. | 2005a | China | SARS-CoV-1 | 28 | 195 | 14·4 (9·8-19·7) |
| Wu et al. | 2005b | China | SARS-CoV-1 | 17 | 131 | 13·0 (7·7-19·3) |
| Yadav et al. | 2021 | India | SARS-CoV-2 | 13 | 100 | 13·0 (7·0-20·4) |
| Zhang et al. | 2020a | China | SARS-CoV-2 | 23 | 30 | 76·7 (59·6-90·3) |
| **Summary Effect** | | | | **895** | **4638** | **19·2 (13·5-25·7)** |
| **Moderate-to-severe anxiety, ongoing** | | | | | | |
| Bonazza et al. | 2020 | Italy | SARS-CoV-2 | 35 | 261 | 13·4 (9·5-17·8) |
| D´Cruz et al. | 2021 | UK | SARS-CoV-2 | 25 | 113 | 22·1 (14·9-30·3) |
| Kwek et al. | 2006 | Singapore | SARS-CoV-1 | 11 | 63 | 17·5 (9·0-27·9) |
| Poyraz et al. | 2021 | Turkey | SARS-CoV-2 | 50 | 284 | 17·6 (13·4-22·3) |
| Raman et al. | 2021 | UK | SARS-CoV-2 | 8 | 57 | 14·0 (6·0-24·4) |
| Rass et al. | 2021 | Austria | SARS-CoV-2 | 7 | 98 | 7·1 (2·7-13·2) |
| van den Borst et al. | 2020 | Netherlands | SARS-CoV-2 | 12 | 124 | 9·7 (5·0-15·6) |
| Wu et al. | 2005b | China | SARS-CoV-1 | 18 | 131 | 13·7 (8·3-20·2) |
| **Summary Effect** | | | | **166** | **1131** | **14·1 (11·1-17·5)** |

**Table A.6:** Continued

| Severity, Domain, Timepoint First author | Year | Country | Epidemic | No. of cases | Sample size | Pooled prevalence (95% CI) |
|---|---|---|---|---|---|---|
| **Moderate-to-severe anxiety, post-illness** | | | | | | |
| Jeong et al. | 2016 | South Korea | MERS-CoV | 7 | 36 | 19·4 (7·9-34·2) |
| Mak et al. | 2009 | China | SARS-CoV-1 | 14 | 90 | 15·6 (8·7-23·9) |
| Park et al. | 2020 | South Korea | MERS-CoV | 9 | 63 | 14·3 (6·6-24·1) |
| Secor et al. | 2020 | Liberia | Ebolavirus | 20 | 203 | 9·9 (6·1-14·4) |
| Secor et al. | 2020 | Sierra Leone | Ebolavirus | 80 | 751 | 10·7 (8·5-13·0) |
| Secor et al. | 2020 | Guinea | Ebolavirus | 22 | 530 | 4·2 (2·6-6·0) |
| **Summary Effect** | | | | **152** | **1673** | **12·1 (5·3-21·1)** |
| **At least mild depression, acute** | | | | | | |
| Akinci and Basar | 2021 | Turkey | SARS-CoV-2 | 82 | 189 | 43·4 (36·4-50·5) |
| Chen et al. | 2020 | China | SARS-CoV-2 | 10 | 31 | 32·3 (16·8-49·9) |
| Guo et al. | 2020 | China | SARS-CoV-2 | 62 | 103 | 60·2 (50·5-69·5) |
| He et al. | 2021 | China | SARS-CoV-2 | 31 | 65 | 47·7 (35·6-59·9) |
| Heyns et al. | 2021 | Belgium | SARS-CoV-2 | 16 | 47 | 34·0 (21·1-48·3) |
| Hu et al. | 2020 | China | SARS-CoV-2 | 39 | 85 | 45·9 (35·4-56·6) |
| Islam et al. | 2021 | Bangladesh | SARS-CoV-2 | 698 | 1002 | 69·7 (66·8-72·5) |
| Ju et al. | 2021 | China | SARS-CoV-2 | 38 | 95 | 40·0 (30·3-50·1) |
| Ju et al. | 2021 | China | SARS-CoV-2 | 29 | 95 | 30·5 (21·6-40·2) |
| Kandeger et al. | 2020 | Turkey | SARS-CoV-2 | 29 | 84 | 34·5 (24·7-45·1) |
| Kang et al. | 2020 | South Korea | SARS-CoV-2 | 26 | 107 | 24·3 (16·6-32·9) |
| Kim et al. | 2018 | South Korea | MERS-CoV | 11 | 27 | 40·7 (22·8-60·0) |
| Kim et al. | 2020 | South Korea | SARS-CoV-2 | 13 | 33 | 39·4 (23·3-56·7) |
| Kong et al. | 2020 | China | SARS-CoV-2 | 41 | 144 | 28·5 (21·4-36·1) |
| Li et al. | 2020 | China | SARS-CoV-2 | 50 | 99 | 50·5 (40·6-60·4) |
| Ma et al. | 2020 | China | SARS-CoV-2 | 332 | 770 | 43·1 (39·6-46·6) |
| Martillo et al. | 2021 | USA | SARS-CoV-2 | 17 | 42 | 40·5 (26·0-55·8) |
| Mina et al. | 2021 | Bangladesh | SARS-CoV-2 | 82 | 145 | 56·6 (48·4-64·5) |
| Park et al. | 2020 | South Korea | MERS-CoV | 50 | 63 | 79·4 (68·4-88·6) |
| Parker et al. | 2021 | USA | SARS-CoV-2 | 17 | 58 | 29·3 (18·2-41·8) |
| Paz et al. | 2020 | Ecuador | SARS-CoV-2 | 399 | 759 | 52·6 (49·0-56·1) |
| Sahan et al. | 2021 | Turkey | SARS-CoV-2 | 118 | 281 | 42·0 (36·3-47·8) |
| Samrah et al. | 2020 | Jordan | SARS-CoV-2 | 29 | 66 | 43·9 (32·1-56·1) |
| Speth et al. | 2020 | Switzerland | SARS-CoV-2 | 54 | 114 | 47·4 (38·2-56·6) |
| Wang et al. | 2021 | China | SARS-CoV-2 | 246 | 460 | 53·5 (48·9-58·0) |
| Yadav et al. | 2021 | India | SARS-CoV-2 | 27 | 100 | 27·0 (18·7-36·2) |
| Zarghami et al. | 2020 | Iran | SARS-CoV-2 | 13 | 30 | 43·3 (26·0-61·5) |
| Zarghami et al. | 2020 | Iran | SARS-CoV-2 | 18 | 52 | 34·6 (22·2-48·2) |
| Zhang et al. | 2020a | China | SARS-CoV-2 | 30 | 30 | 100·0 (94·3-100·0) |
| Zhang et al. | 2020b | China | SARS-CoV-2 | 55 | 296 | 18·6 (14·3-23·2) |
| **Summary Effect** | | | | **2662** | **5472** | **45·3 (38·0-52·7)** |

**Table A.6:** Continued

| Severity, Domain, Timepoint<br>First author | Year | Country | Epidemic | No. of cases | Sample size | Pooled prevalence (95% CI) |
|---|---|---|---|---|---|---|
| **At least mild depression, ongoing** | | | | | | |
| Bonazza et al. | 2020 | Italy | SARS-CoV-2 | 44 | 261 | 16·9 (12·5-21·7) |
| Kwek et al. | 2006 | Singapore | SARS-CoV-1 | 17 | 63 | 27·0 (16·7-38·7) |
| Olanipekun et al. | 2021 | USA | SARS-CoV-2 | 51 | 73 | 69·9 (58·8-79·9) |
| Raman et al. | 2021 | UK | SARS-CoV-2 | 24 | 57 | 42·1 (29·5-55·2) |
| Rass et al. | 2021 | Austria | SARS-CoV-2 | 8 | 98 | 8·2 (3·4-14·5) |
| **Summary Effect** | | | | **144** | **552** | **30·8 (11·7-54·1)** |
| **At least mild depression, post-illness** | | | | | | |
| Bah et al. | 2020 | Sierra Leone | Ebolavirus | 93 | 197 | 47·2 (40·3-54·2) |
| Luyt et al. | 2012 | France | H1N1 | 10 | 37 | 27·0 (13·8-42·6) |
| Park et al. | 2020 | South Korea | MERS-CoV | 35 | 63 | 55·6 (43·1-67·7) |
| Secor et al. | 2020 | Liberia | Ebolavirus | 96 | 198 | 48·5 (41·5-55·5) |
| Secor et al. | 2020 | Sierra Leone | Ebolavirus | 350 | 751 | 46·6 (43·0-50·2) |
| Secor et al. | 2020 | Guinea | Ebolavirus | 178 | 529 | 33·6 (29·7-37·7) |
| **Summary Effect** | | | | **762** | **1775** | **43·5 (34·1-53·1)** |
| **Moderate-to-severe depression, acute** | | | | | | |
| Chen et al.. | 2021 | China | SARS-CoV-2 | 189 | 898 | 21·0 (18·4-23·8) |
| Chen et al. | 2020 | China | SARS-CoV-2 | 2 | 31 | 6·5 (0·1-18·5) |
| Guo et al. | 2020 | China | SARS-CoV-2 | 18 | 103 | 17·5 (10·7-25·5) |
| He et al. | 2021 | China | SARS-CoV-2 | 18 | 65 | 27·7 (17·4-39·3) |
| Hu et al. | 2020 | China | SARS-CoV-2 | 21 | 85 | 24·7 (16·1-34·5) |
| Islam et al. | 2021 | Bangladesh | SARS-CoV-2 | 482 | 1002 | 48·1 (45·0-51·2) |
| Jeong et al. | 2020 | South Korea | SARS-CoV-2 | 20 | 126 | 15·9 (10·0-22·8) |
| Ju et al. | 2021 | China | SARS-CoV-2 | 17 | 95 | 17·9 (10·8-26·3) |
| Ju et al. | 2021 | China | SARS-CoV-2 | 13 | 95 | 13·7 (7·4-21·4) |
| Kang et al. | 2020 | South Korea | SARS-CoV-2 | 7 | 107 | 6·5 (2·5-12·1) |
| Kim et al. | 2018 | South Korea | MERS-CoV | 3 | 27 | 11·1 (1·5-26·3) |
| Kong et al. | 2020 | China | SARS-CoV-2 | 21 | 144 | 14·6 (9·2-20·9) |
| Li et al. | 2020 | China | SARS-CoV-2 | 29 | 99 | 29·3 (20·7-38·7) |
| Ma et al. | 2020 | China | SARS-CoV-2 | 143 | 770 | 18·6 (15·9-21·4) |
| Martillo et al. | 2021 | USA | SARS-CoV-2 | 8 | 42 | 19·0 (8·4-32·5) |
| Park et al. | 2020 | South Korea | MERS-CoV | 42 | 63 | 66·7 (54·5-77·8) |
| Parker et al. | 2021 | USA | SARS-CoV-2 | 12 | 58 | 20·7 (11·1-32·2) |
| Paz et al. | 2020 | Ecuador | SARS-CoV-2 | 154 | 759 | 20·3 (17·5-23·2) |
| Samrah et al. | 2020 | Jordan | SARS-CoV-2 | 14 | 66 | 21·2 (12·1-32·0) |
| Speth et al. | 2020 | Switzerland | SARS-CoV-2 | 24 | 114 | 21·1 (14·0-29·1) |
| Wang et al. | 2021 | China | SARS-CoV-2 | 114 | 460 | 24·8 (20·9-28·8) |
| Wu et al. | 2005a | China | SARS-CoV-1 | 35 | 195 | 17·9 (12·9-23·7) |
| Wu et al. | 2005b | China | SARS-CoV-1 | 23 | 131 | 17·6 (11·5-24·6) |
| Xu et al. | 2021 | China | SARS-CoV-2 | 12 | 121 | 9·9 (5·1-16·0) |
| Yadav et al. | 2021 | India | SARS-CoV-2 | 6 | 100 | 6·0 (2·0-11·6) |
| Zhang et al. | 2020a | China | SARS-CoV-2 | 23 | 30 | 76·7 (59·6-90·3) |
| **Summary Effect** | | | | **1450** | **5786** | **21·9 (15·8-28·6)** |

**Table A.6:** Continued

| Severity, Domain, Timepoint<br>First author | Year | Country | Epidemic | No. of cases | Sample size | Pooled prevalence (95% CI) |
|---|---|---|---|---|---|---|
| **Moderate-to-severe depression, ongoing** | | | | | | |
| Bonazza et al. | 2020 | Italy | SARS-CoV-2 | 27 | 261 | 10·3 (6·9-14·4) |
| D´Cruz et al. | 2021 | UK | SARS-CoV-2 | 20 | 111 | 18·0 (11·4-25·8) |
| Kwek et al. | 2006 | Singapore | SARS-CoV-1 | 7 | 63 | 11·1 (4·3-20·2) |
| Olanipekun et al. | 2021 | USA | SARS-CoV-2 | 40 | 73 | 54·8 (43·2-66·1) |
| Poyraz et al. | 2021 | Turkey | SARS-CoV-2 | 51 | 284 | 18·0 (13·7-22·7) |
| Raman et al. | 2021 | UK | SARS-CoV-2 | 11 | 57 | 19·3 (10·0-30·7) |
| Rass et al. | 2021 | Austria | SARS-CoV-2 | 3 | 98 | 3·1 (0·4-7·6) |
| van den Borst et al. | 2020 | Netherlands | SARS-CoV-2 | 14 | 124 | 11·3 (6·2-17·5) |
| Wu et al. | 2005b | China | SARS-CoV-1 | 17 | 131 | 13·0 (7·7-19·3) |
| **Summary Effect** | | | | **190** | **1202** | **16·0 (8·6-25·2)** |
| **Moderate-to-severe depression, post-illness** | | | | | | |
| Etard et al. | 2017 | Guinea | Ebolavirus | 97 | 472 | 20·6 (17·0-24·3) |
| Keita et al. | 2017 | Guinea | Ebolavirus | 38 | 256 | 14·8 (10·7-19·5) |
| Lee et al. | 2019 | South Korea | MERS-CoV | 14 | 52 | 26·9 (15·6-39·9) |
| Lee et al. | 2019 | South Korea | MERS-CoV | 9 | 52 | 17·3 (8·1-28·9) |
| Mak et al. | 2009 | China | SARS-CoV-1 | 17 | 90 | 18·9 (11·4-27·7) |
| Park et al. | 2020 | South Korea | MERS-CoV | 17 | 63 | 27·0 (16·7-38·7) |
| Secor et al. | 2020 | Liberia | Ebolavirus | 43 | 198 | 21·7 (16·2-27·8) |
| Secor et al. | 2020 | Sierra Leone | Ebolavirus | 136 | 751 | 18·1 (15·4-20·9) |
| Secor et al. | 2020 | Guinea | Ebolavirus | 49 | 529 | 9·3 (6·9-11·9) |
| **Summary Effect** | | | | **420** | **2463** | **18·2 (13·8-23·0)** |
| **At least mild post-traumatic stress, acute** | | | | | | |
| Bellan et al. | 2021 | Italy | SARS-CoV-2 | 102 | 238 | 42·9 (36·6-49·2) |
| Chieffo et al. | 2020 | Italy | SARS-CoV-2 | 27 | 33 | 81·8 (66·6-93·4) |
| **Summary Effect** | | | | **129** | **271** | **62·4 (23·6-93·8)** |
| **At leat mild post-traumatic stress, ongoing** | | | | | | |
| Kwek et al. | 2006 | Singapore | SARS-CoV-1 | 26 | 63 | 41·3 (29·4-53·7) |
| Poyraz et al. | 2021 | Turkey | SARS-CoV-2 | 124 | 284 | 43·7 (37·9-49·5) |
| **Summary Effect** | | | | **150** | **347** | **43·2 (38·0-48·5)** |
| **At least mild post-traumatic stress, post-illness** | | | | | | |
| Chieffo et al. | 2020 | Italy | SARS-CoV-2 | 7 | 14 | 50·0 (23·7-76·3) |
| Lee et al. | 2019 | South Korea | MERS-CoV | 22 | 52 | 42·3 (29·1-56·0) |
| Lee et al. | 2019 | South Korea | MERS-CoV | 14 | 52 | 26·9 (15·6-39·9) |
| Park et al. | 2020 | South Korea | MERS-CoV | 27 | 63 | 42·9 (30·8-55·3) |
| **Summary Effect** | | | | **70** | **181** | **38·6 (24·0-54·3)** |

**Table A.6:** Continued

| Severity, Domain, Timepoint<br>First author | Year | Country | Epidemic | No. of cases | Sample size | Pooled prevalence (95% CI) |
|---|---|---|---|---|---|---|
| **Moderate-to-severe post-traumatic stress, acute** | | | | | | |
| Bellan et al. | 2021 | Italy | SARS-CoV-2 | 41 | 238 | 17·2 (12·7-22·3) |
| Chieffo et al. | 2020 | Italy | SARS-CoV-2 | 19 | 33 | 57·6 (40·2-74·1) |
| Horn et al. | 2020 | France | SARS-CoV-2 | 60 | 179 | 33·5 (26·8-40·6) |
| Mazza et al. | 2020 | Italy | SARS-CoV-2 | 71 | 300 | 23·7 (19·0-28·7) |
| Mazza et al. | 2020 | Italy | SARS-CoV-2 | 34 | 102 | 33·3 (24·5-42·8) |
| Wu et al. | 2005a | China | SARS-CoV-1 | 11 | 195 | 5·6 (2·8-9·4) |
| Wu et al. | 2005b | China | SARS-CoV-1 | 6 | 131 | 4·6 (1·5-8·9) |
| **Summary Effect** | | | | **242** | **1178** | **25·3 (7·5-48·9)** |
| **Moderate-to-severe post-traumatic stress, ongoing** | | | | | | |
| Bonazza et al. | 2020 | Italy | SARS-CoV-2 | 67 | 184 | 36·4 (29·6-43·5) |
| Kwek et al. | 2006 | Singapore | SARS-CoV-1 | 23 | 63 | 36·5 (25·0-48·8) |
| Poyraz et al. | 2021 | Turkey | SARS-CoV-2 | 72 | 284 | 25·4 (20·5-30·6) |
| van den Borst et al. | 2020 | Netherlands | SARS-CoV-2 | 12 | 122 | 9·8 (5·1-15·8) |
| Wu et al. | 2005b | China | SARS-CoV-1 | 7 | 131 | 5·3 (2·0-10·0) |
| **Summary Effect** | | | | **181** | **784** | **20·9 (9·2-35·7)** |
| **Moderate-to-severe post-traumatic stress, post-illness** | | | | | | |
| Chieffo et al. | 2020 | Italy | SARS-CoV-2 | 4 | 14 | 28·6 (7·4-55·4) |
| Luyt et al. | 2012 | France | H1N1 | 16 | 37 | 43·2 (27·6-59·6) |
| **Summary Effect** | | | | **20** | **51** | **39·1 (25·7-53·2)** |
| **Moderate-to-severe general distress, ongoing** | | | | | | |
| Cheng et al. | 2004 | China | SARS-CoV-1 | 68 | 100 | 68·0 (58·5-76·8) |
| Sheng et al. | 2005 | China | SARS-CoV-1 | 66 | 102 | 64·7 (55·1-73·7) |
| **Summary Effect** | | | | **134** | **202** | **66·3 (59·6-72·7)** |
| **Single prevalence rates (no pooling)** | | | | | | |
| Wang et al.<br>(mild somatization, acute) | 2021 | China | SARS-CoV-2 | 304 | 460 | 66·1 (61·7-70·3) |
| Mowla et al.<br>(moderate general distress, acute) | 2021 | Iran | SARS-CoV-2 | 69 | 69 | 100·0 (97·5-100·0) |
| Wang et al.<br>(moderate somatization, acute) | 2021 | China | SARS-CoV-2 | 167 | 460 | 36·3 (32·0-40·8) |
| Lam et al.<br>(depression and anxiety jointly, post-illness) | 2009 | China | SARS-CoV-1 | 83 | 170 | 48·8 (41·3-56·4) |
| Lee et al.<br>(moderate general distress, post-illness) | 2007 | China | SARS-CoV-1 | 61 | 96 | 63·5 (53·6-72·9) |

**Table A.7:** Test of moderators on at least mild or moderate-to-severe mental health problems including all domains. All moderators were tested separately adjusted for follow-up time in months.

| | At least mild mental health problems | | Moderate-to-severe mental health problems | |
|---|---|---|---|---|
| **Moderator variable** | **Coefficient$_{arcsine}$ (95% CI)** | ***p*-value** | **Coefficient$_{arcsine}$ (95% CI)** | ***p*-value** |
| Proportion of females | 0·001 (-0·005 to 0·006) | *0·7390* | <0·000 (-0·005 to 0·004) | *0·8835* |
| Mean/median (years) | -0·007 (-0·014 to 0·001) | *0·0926* | -0·001 (-0·007 to 0·005) | *0·7696* |
| Proportion with higher education | -0·004 (-0·008 to 0·001) | *0·1023* | -0·002 (-0·006 to 0·003) | *0·4891* |
| Proportion with previous mental health condition | -0·005 (-0·016 to 0·006) | *0·3292* | 0·003 (-0·006 to 0·013) | *0·5027* |
| Proportion of health care workers | -0·007 (-0·019 to 0·005) | *0·2425* | -0·004 (-0·014 to 0·006) | *0·4457* |
| Duration of hospitalization (days) | 0·010 (-0·007 to 0·026) | *0·2427* | 0·007 (-0·004 to 0·018) | *0·2066* |
| Proportion with intensive care unit treatment | 0·000 (-0·004 to 0·004) | *0·9523* | 0·002 (-0·001 to 0·005) | *0·1456* |
| Type of treatment | | | | |
| Inpatient treatment | reference | | reference | |
| Miscellaneous | -0·022 (-0·232 to 0·188) | *0·8349* | -0·072 (-0·217 to 0·072) | *0·3227* |
| Outpatient | -0·018 (-0·252 to 0·217) | *0·8821* | 0·021 (-0·174 to 0·216) | *0·8301* |
| Response rate (%) | 0·000 (-0·003 to 0·002) | *0·8243* | 0·000 (-0·002 to 0·003) | *0·8859* |
| Sampling | | | | |
| Non-random | reference | | reference | |
| Random | -0·161 (-0·280 to-0·043) | *0·0083* | -0·015 (-0·095 to 0·065) | *0·7122* |
| World region | | | | |
| China | reference | | reference | |
| Other Asia | -0·066 (-0·235 to 0·103) | *0·4366* | 0·023 (-0·110 to 0·157) | *0·7284* |
| Europe | -0·112 (-0·276 to 0·051) | *0·1753* | 0·013 (-0·110 to 0·137) | *0·8309* |
| Africa | 0·047 (-0·296 to 0·390) | *0·7869* | 0·046 (-0·188 to 0·281) | *0·6955* |
| America | -0·007 (-0·246 to 0·232) | *0·9531* | 0·068 (-0·126 to 0·262) | *0·4905* |
| Epidemic | | | | |
| SARS-CoV-2 | reference | | reference | |
| SARS-CoV-1 | 0·231 (-0·146 to 0·608) | *0·2260* | 0·067 (-0·141 to 0·275) | *0·5252* |
| MERS-CoV | 0·102 (-0·169 to 0·372) | *0·4568* | 0·145 (-0·038 to 0·329) | *0·1183* |
| Ebolavirus | 0·229 (-0·158 to 0·616) | *0·2424* | 0·113 (-0·123 to 0·349) | *0·3458* |
| H1N1 | 0·116 (-0·336 to 0·568) | *0·6097* | 0·392 (0·003 to 0·782) | *0·0483* |
| Timepoint of follow-up | | | | |
| Acute | Reference | | Reference | |
| Mild | -0·130 (-0·305 to 0·045) | *0·1432* | -0·044 (-0·135 to 0·047) | *0·3379* |
| Post-illness | -0·184 (-0·307 to -0·061) | *0·0038* | -0·146 (-0·253 to -0·039) | *0·0080* |

Shown are coefficients for a meta-regression model including arcsine transformed proportions. In categorical moderators the coefficients show the comparison between a respective factor level and the reference group. H1N1=Swine flu. MERS-CoV=Middle East respiratory syndrome coronavirus. SARS-CoV-1=Severe acute respiratory syndrome coronavirus 1. SARS-CoV-2=Severe acute respiratory syndrome coronavirus 2. All moderators were tested separately adjusted for follow-up time in month. A moderator analysis including all moderators jointly was not possible due to the substantial number of missing information across the studies.
